# Type 1 Diabetes Polygenic Scores Improve Diagnostic Accuracy in Pediatric Diabetes Care

**DOI:** 10.1101/2025.03.06.25323229

**Authors:** Raymond J. Kreienkamp, Aaron J. Deutsch, Alicia Huerta-Chagoya, Erin M. Borglund, Jose C. Florez, Jason Flannick, Miriam S. Udler

**Affiliations:** Division of Endocrinology, Department of Pediatrics, Boston Children’s Hospital, Boston, MA, USA; Center for Genomic Medicine and Diabetes Unit, Endocrine Division, Department of Medicine, Massachusetts General Hospital, Boston, MA, USA; Department of Medicine, Harvard Medical School, Boston, MA, USA; Programs in Metabolism and Medical and Population Genetics, The Broad Institute of MIT and Harvard, Cambridge, MA, USA; Computational Health Informatics Program (CHIP), Boston Children’s Hospital, Boston, MA, USA; Division of Genetics and Genomics, Department of Pediatrics, Boston Children’s Hospital, Boston, MA, USA

**Author notes:** Corresponding Author: Raymond J. Kreienkamp, MD, PhD.

**Keywords:** pediatric diabetes, genetics, polygenic scores

## Abstract

**Background:** Accurately classifying pediatric diabetes can be challenging for providers, and misclassification can result in suboptimal care. In recent years, type 1 diabetes (T1D) polygenic scores, which quantify one’s genetic risk for T1D based on T1D risk allele burden, have been developed with good discriminating capacity between T1D and not-T1D. These tools have the potential to improve significantly diagnostic provider accuracy if used in clinic.

**Methods:** We applied T1D polygenic scores to a group of pediatric patients (n=1846) with genetic data available in the Boston Children’s Hospital PrecisionLink Biobank, including 96 individuals diagnosed with T1D.

**Results:** Patients with a clinical diagnosis of T1D had higher T1D polygenic scores compared to controls (Wilcoxon rank-sum *P*<0.0001). Sixty-nine of the 74 individuals with diabetes and a T1D polygenic score exceeding an externally validated cutoff for distinguishing T1D from not-T1D were confirmed to have T1D. There were multiple cases where T1D polygenic scores would have clinical utility. An elevated T1D polygenic score suggested T1D in a pancreatic autoantibody (PAA)-negative individual with negative MODY genetic testing and a phenotype matching T1D. A low T1D polygenic score accurately indicated atypical diabetes in an individual found to have *HNF1B*-MODY. One individual had positive PAA, but the provider noted that the patient may not have classic T1D, as later suggested by a low T1D polygenic score.

**Conclusion:** T1D polygenic scores already have clinical utility to aid in the accurate diagnosis of pediatric diabetes. Efforts are now needed to advance their use in clinical practice.

## Introduction

Accurately diagnosing pediatric diabetes is critical for optimal clinical care but is often delayed or incorrect. Inaccurate diagnoses occur because current differentiating factors cannot accurately distinguish between type 1 diabetes (T1D) and other etiologies in all cases. Pancreatic autoantibodies (PAA) can be present in both T1D, where they are more common, and type 2 diabetes (T2D) [1-4]. Obesity is a known risk factor for T2D [5, 6], but 25%-40% of youth with T1D are overweight or obese [7, 6]. DKA, while traditionally associated with T1D, can also be seen in youth with T2D [8]. Maturity-onset diabetes of the young (MODY) can present similarly to both T1D and T2D, causing greater than 80% of MODY to be missed and incorrectly classified [9]. Missed or inaccurate diagnoses are becoming more consequential, as misdiagnosis could delay the use of emerging T1D preventive therapies [10-14], direct inappropriate initiation or delay in insulin therapy [15, 16], or prevent the use of targeted therapies in MODY [17]. Therefore, new tools are needed to improve pediatric diabetes diagnosis and care.

T1D polygenic scores estimate an individual’s genetic liability for T1D based on the aggregate count of the number and effect size of risk variants present. While multiple T1D polygenic scores have been developed, two scores are most utilized in research studies. Sharp *et al*. developed GRS2 in a European ancestry population (score hereafter called T1D_GRS_EUR), which used 67 single nucleotide polymorphisms (SNPs) accounting for interactions between 18 HLA DR-DQ haplotype combinations, and was found to be highly discriminative for all T1D (area under the receiver operating characteristic curve [AUC] 0.92) [18]. While initially developed in a European ancestry population, this score might also be discriminatory in other ancestries [19], though different score thresholds may be needed to indicate an equivalent risk. Onengut-Gumuscu *et al*. developed an African ancestry-specific T1D polygenic score (hereafter called T1D_GRS_AFR) with 7 SNPs with strong prediction of T1D risk (AUC 0.871) [20]. T1D polygenic scores have demonstrated utility to differentiate T1D from other forms of diabetes, including T2D (AUC 0.88) [21], MODY (AUC 0.87) [22], and, combined with age of diagnosis, monogenic autoimmunity (AUC 0.88) [23].

Given the high predictive power of T1D polygenic scores in a research setting, these tools have the potential to improve clinical care in a real-world clinical setting. To test their clinical utility and identify specific scenarios where polygenic scores could be helpful in clinical practice now, we applied T1D polygenic scores to a group of pediatric patients in the Boston Children’s Hospital PrecisionLink Biobank for Health Discovery (PLB) [24], which includes genetic data and corresponding clinical information.

## Methods

### PrecisionLink Biobank and Genetic Data

Genetic data was obtained from the PLB, a hospital-sponsored centralized source of retrospectively consented biospecimens, genomic, and phenotype data supporting broad research use [24]. All Boston Children’s Hospital (BCH) patients are eligible for enrollment. Written informed consent was obtained for all participants which provided permission for (1) research use of electronic health record (EHR) data including demographics, dates and types of encounters (e.g., ED visit, hospitalization), clinic notes, diagnoses, laboratory values, medications prescribed, and procedures performed; (2) broad research use of residual specimens produced as by-products of clinical care; and (3) an optional additional collection of a research tube during a future clinical laboratory draw.

PLB specimens were processed and stored in the Biobank Core Lab facility at BCH. DNA was then extracted from clinical specimens and sent for genotyping. Genetic data was then imputed with TOPMed Imputation Server using TOPMed reference panel r2 [25, 26]. The imputation server used the minimac4 (1.5.7) algorithm for genotype imputation (with Eagle for phasing) [27]. Only autosomal variants were used for imputation.

The PLB initiative is approved by the BCH Institutional Review Board (protocol number - P00000159). This project (Genetic and clinical features of diabetes and metabolic conditions) is approved by the BCH Institutional Review Board (protocol number - P00044203).

### Clinical Characteristics and Diabetes Classification

Individuals were classified as having diabetes or not diabetes by ICD9 codes 250 or ICD10 codes E8-E13. If either of these were present, the individual was classified as having diabetes. If these were not present, the individual was classified as a control. All individuals classified as having diabetes were then manually reviewed by a pediatric endocrinology fellow to confirm the diagnosis. Classification was refined from the ICD code to that diagnosis most consistent with the clinical phenotype by review of provider notes, genetic testing, and clinical course at that time. Not all individuals followed at BCH for their diabetes care, so the amount of available data (including PAA) was variable throughout the cohort. Additionally, these individuals were diagnosed with diabetes over a time span of 25 years, so clinical practice evolved with regards to the labs that are obtained with new-onset diabetes (e.g. number of PAA screened, if at all screened). Individuals were classified as having T1D, T2D, MODY, cystic-fibrosis related diabetes (CFRD), steroid-induced hyperglycemia (SIH), diabetes coded in error, or other diabetes type (including secondary to cancer therapy, pancreatitis, etc.). Given the desire to focus on pediatric diabetes, those with diabetes diagnosis after age 21 were excluded. Genetic ancestry was determined for individuals in PLB, using techniques previously utilized [28] in Mass General Brigham Biobank (MGBB).

### Polygenic Score Generation

T1D polygenic scores were computed from available imputed genetic data, as previously described [20, 18]. Genetic ancestry was used to select the appropriate T1D polygenic score. T1D_GRS_EUR was computed in all individuals. T1D_GRS_EUR score percentiles were noted as previously reported in individuals with T1D in the UK Biobank (UKBB) European population [18]. T1D_GRS_AFR was computed in African ancestry individuals. Two SNPs in the original T1D_GRS_AFR were not available in PLB and were replaced with SNPs in linkage disequilibrium with the original SNPs. The original SNP rs34303755 was replaced with rs34036879, and rs2187668 was replaced with rs1617322. T1D_GRS_AFR score percentiles were reported as found in African ancestry individuals with T1D, as verified by manual curation [29], in MGBB [30].

### Statistics

Polygenic score distributions are presented as boxplots, with the median and range extending to 1.5x interquartile distance or end of range, whichever occurs first. For T1D_GRS_EUR boxplot figures, a line was drawn at 12.88, the point previously reported as the optimal cutoff based on the highest Youden index for separating T1D cases from controls [18]. A line was drawn at 4.4815 in T1D_GRS_AFR as the point with the highest Youden index as found in MGBB to discriminate T1D from not-T1D. Distribution medians were compared using Wilcoxon rank-sum tests, with Bonferroni correction when needed. All plots, including receiver operating characteristic curves, were generated in R.

## Results

### Overall Cohort Characteristics

We reviewed clinical records for individuals with genetic data available in PLB. There were 147 individuals with diabetes mellitus or hyperglycemia, of whom 96 had a clinical diagnosis of T1D, and the remaining individuals had T2D, CFRD, SIH, or other (Table 1). The participants with diabetes were predominantly European ancestry (82%) and with T1D (65%). Analyses were focused on individuals with T1D and of European ancestry given lower sample sizes among other diagnoses and ancestries.

**Table 1.** Clinician-reviewed diagnoses of individuals with diabetes or hyperglycemia in Boston Children’s Hospital PrecisionLink Biobank.

|  | Overall (n) | European<br>Ancestry (n) | Other<br>Ancestry(n) |
| --- | --- | --- | --- |
| Total | 1846 | 1361 | 485 |
| Controls (Individuals without Diabetes) | 1699 | 1240 | 459 |
| Total Diabetes | 147 | 121 | 26 |
| Type 1 Diabetes | 96 | 83 | 13 |
| Type 2 Diabetes | 13 | 4 | 9 |
| Cystic Fibrosis-Related Diabetes | 20 | 18 | 2 |
| Steroid-Induced Hyperglycemia | 8 | 8 | 0 |
| Other Diabetes Type | 10 | 8 | 2 |

T1D_GRS_EUR scores were significantly different between controls and T1D (*P* < 0.0001) (Figure 1A). In the PLB European ancestry cohort, individuals with T1D were the only group with a T1D polygenic score that was significantly different from that seen in controls (*P*=1 for comparisons between controls and other diabetes types) (Figure 1B). The T1D_GRS_EUR had good differentiating capacity between T1D and not-T1D in PLB overall (AUC=0.89) as well as in European ancestry individuals (AUC=0.90) (Figure 2).

**Figure 1.**
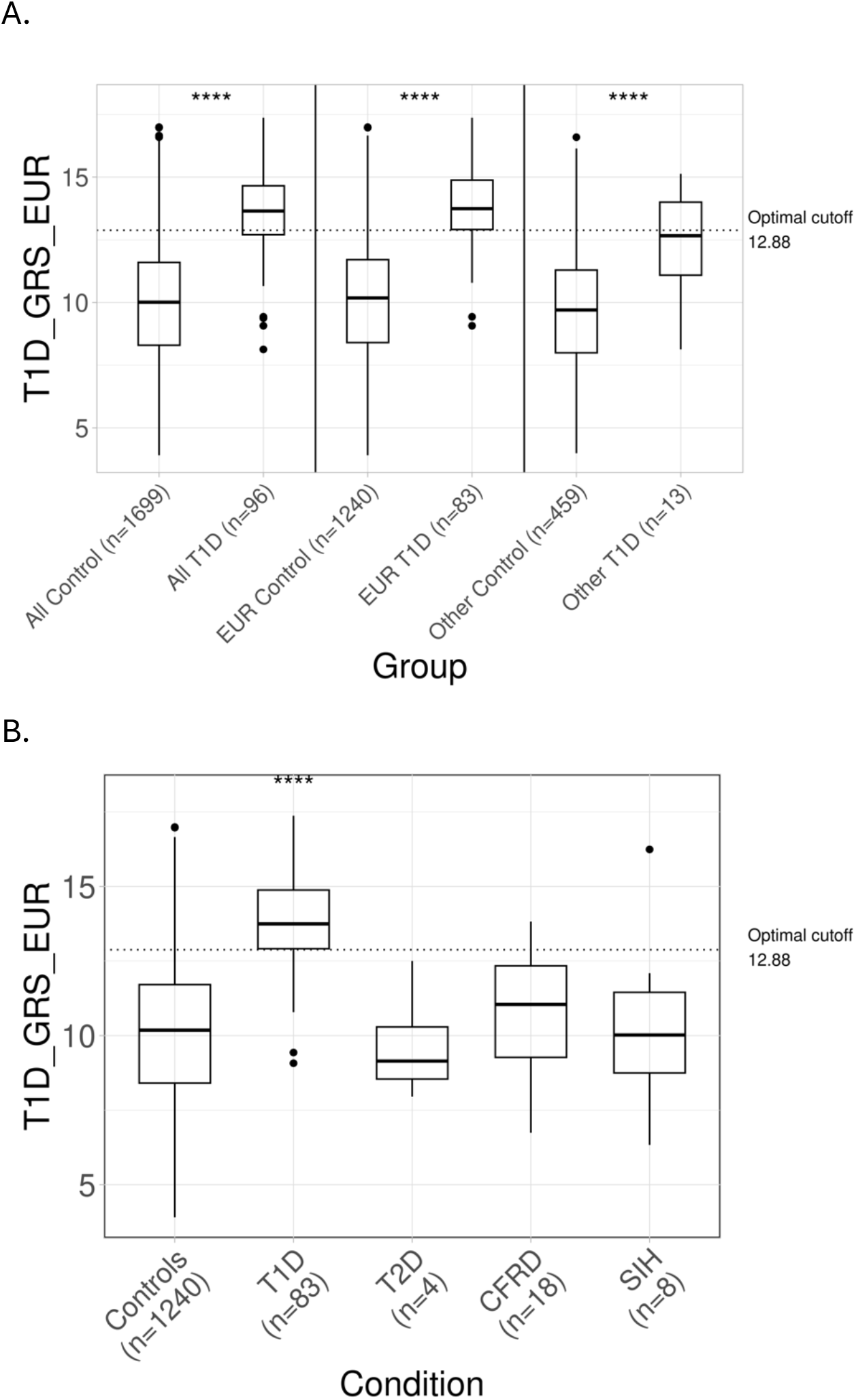
T1D Polygenic Score Performance in Boston Children’s Hospital PrecisionLink Biobank (PLB). (A) T1D_GRS_EUR performance in PLB for all individuals, European ancestry individuals, and non-European (Other) ancestry individuals, without diabetes or with T1D. Dotted line is 12.88, the optimal point for distinguishing T1D from not-T1D. (B) T1D_GRS_EUR performance in European PLB in controls, T1D, T2D, CFRD, and SIH. Dotted line is 12.88, the optimal point for distinguishing T1D from not-T1D. T1D, type 1 diabetes; T2D, type 2 diabetes; CFRD, cystic fibrosis-related diabetes; SIH, steroid-induced hyperglycemia; ****, Wilcoxon rank-sum *P*<0.0001.

**Figure 2.**
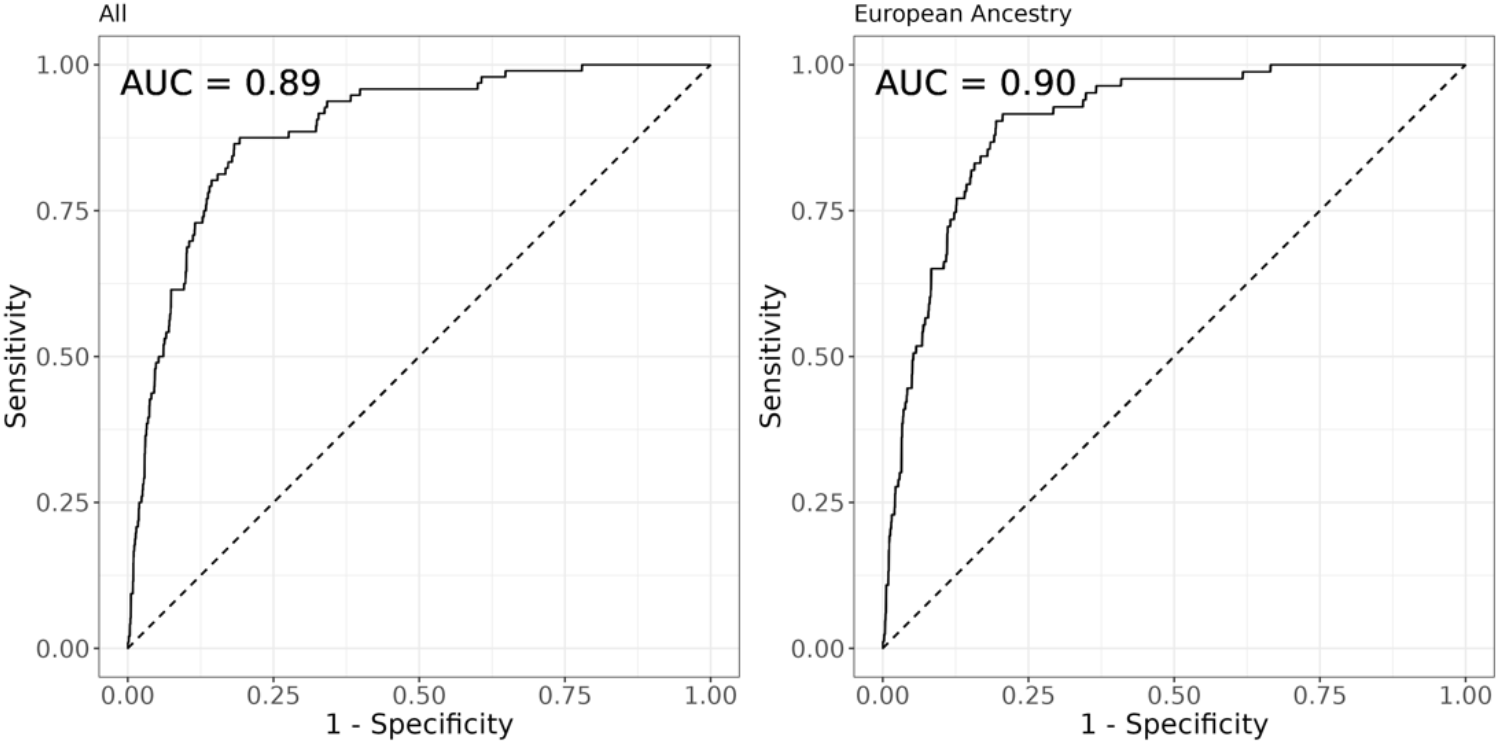
Receiver Operating Characteristic Curves for T1D Polygenic Scores. Receiver operating characteristic curves for T1D_GRS_EUR for differentiating control and T1D in all individuals and European ancestry individuals.

Sixty-nine of 74 individuals with diabetes and a T1D polygenic score exceeding the point with the highest Youden index (T1D_GRS_EUR=12.88), or optimal point for distinguishing T1D from not-T1D, had T1D, resulting in a positive predictive value of 93%. This included 63 of 67 European ancestry individuals (PPV 94%) and 6 of 7 non-European ancestry individuals. Of those 5 individuals with scores above the optimal cutoff without a diagnosis of T1D, there was one individual with CFRD, two individuals with T2D, one individual with SIH, and one individual with diabetes in association with mitochondrial encephalomyopathy, lactic acidosis, and stroke-like episodes (MELAS). Two of these five individuals had hypothyroidism. Three of the five individuals were on chronic insulin therapy.

One case was particularly interesting. While completing these analyses, one teenager classified as having SIH developed a hemoglobin A1c that up-trended to 6.9% while not receiving steroids for over one year. Her BMI was 23.7 kg/m2. She had previously been GAD-65 antibody positive. IA-2 and ZnT8 autoantibodies were checked multiple times, including with up-trending hemoglobin A1c, and were negative. She had a family history of T1D in multiple relatives. Despite the original clinician-based classification of SIH (which has been retained in our reporting results), this individual developed T1D. This diagnosis is supported by BMI, GAD-65 autoantibody positivity, and this T1D polygenic score.

There is increasing likelihood for monogenic autoimmunity with decreasing T1D polygenic score [23]. Seven European ancestry individuals with a clinical diagnosis of T1D had T1D polygenic scores below the T1D 5^th^ percentile (T1D_GRS_EUR 11.68). In our cohort, two individuals had multiple autoimmune conditions. One individual (T1D_GRS_EUR score 11.0) had T1D, Hashimoto’s thyroiditis, celiac disease, and positive 21-hydroxylase autoantibodies. One individual (T1D_GRS_EUR score 10.8) had T1D, hypothyroidism, and celiac disease. None had documentation noting concern for monogenic autoimmunity.

### Autoantibody-negative type 1 diabetes

One proposed benefit of the T1D polygenic score is to identify individuals without PAA positivity, that likely still have T1D. In PLB, there were two European ancestry individuals with a diagnosis of T1D that had negative PAA. One of these individuals had negative PAA and negative MODY genetic testing (Figure 3A). This European ancestry prepubertal girl presented with new-onset diabetes with nocturnal enuresis, polyuria, fatigue, weight loss, and polydipsia. At that time, she was not in DKA, but she had a serum blood glucose 270 mg/dL and hemoglobin A1c 9.9%. Her BMI was at the 23^rd^ percentile. Her family history was significant for T2D and multiple relatives with Graves’ disease. Over one year after diagnosis, she was receiving less insulin than expected, with little hypoglycemia, and her hemoglobin A1c was 6.4%. Monogenic diabetes genetic testing was pursued with a 13-gene panel through the University of Chicago and was negative. The provider later wrote, “Although I recently suspected possible monogenic diabetes due to her lack of antibodies and low insulin requirement, her total daily dose of insulin has increased significantly over the last few months, so in retrospect it is less surprising that her genetic testing was negative.” T1D_GRS_EUR was 15.9, which is in the >75^th^ percentile for those with T1D in UKBB, and above the point with the highest Youden index for distinguishing T1D from not-T1D. Consequently, this result, had the provider been aware, could have reassured the provider that her diagnosis was most likely T1D. The other individual with PAA-negative T1D had a polygenic score 16.1, also above the point with the highest Youden index, also suggesting most likely T1D. She also remained on insulin after years of therapy and followed a typical T1D course.

**Figure 3.**
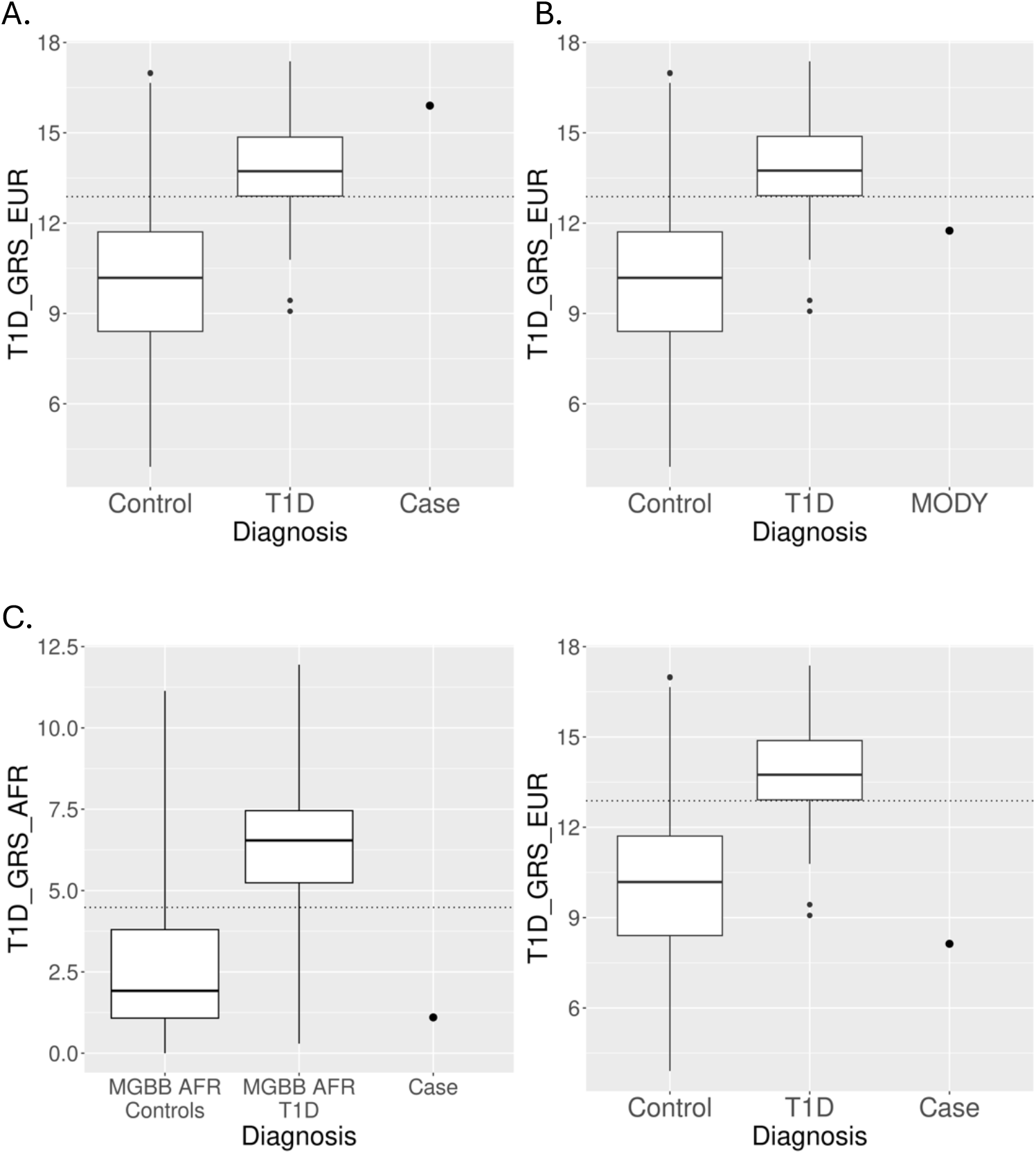
Cases with Improved Diagnostic Accuracy with T1D Polygenic Score. (A) T1D_GRS_EUR score for a European ancestry individual with autoantibody-negative T1D compared to PLB scores for European ancestry controls and individuals with T1D. Note case has score greater than point with highest Youden index and more consistent with T1D. (B) T1D_GRS_EUR score for a European ancestry individual with MODY compared to PLB scores for European ancestry controls and individuals with T1D. Note score is significantly lower than point with highest Youden index, suggestive of not-T1D. (C) T1D_GRS_AFR and T1D_GRS_EUR score for African ancestry individual with an unclear diabetes diagnosis. Score is less than point with highest Youden index in both scores, suggesting not-T1D.

### MODY

One European ancestry teenage female in the cohort was diagnosed with MODY (Figure 3B). She initially presented with polyuria and polydipsia. Blood glucose at presentation was 450 mg/dL, and her hemoglobin A1c was 12.1%. She did not have DKA. She had a three-generation family history of diabetes. There was no known family history of autoimmunity. She was overweight with a BMI at the 92^nd^ percentile. On her initial labs, her magnesium was low at 1.2 mg/dL, so nephrology was consulted. To evaluate her hypomagnesemia and new-onset diabetes in the setting of negative PAAs, genetic testing was obtained, and this demonstrated a 17q12 microdeletion which included the MODY gene *HNF1B*. As a result, she was diagnosed with *HNF1B*-MODY. She had a T1D_GRS_EUR score of 11.7, which is below the point with the highest Youden index for distinguishing T1D from not-T1D. Therefore, her T1D score would have raised suspicion for not-T1D, had it been known to her provider.

### Unclear diabetes diagnosis

One prepubertal African ancestry individual presented with new-onset “T1D” with a hemoglobin A1c 6.7% (Figure 3C). Her BMI was greater than the 99^th^ percentile. She had relatives with diagnoses of T1D or T2D. She had positive GAD-65 and IA-2 autoantibodies. She was assumed to have T1D. Because of this, she was started on insulin. However, she self-discontinued insulin after her hemoglobin A1c fell to 5.8%. Over five years after diagnosis, she did not require insulin and yet had no episodes of DKA. The provider most recently noted that “she is producing endogenous insulin and may have [an alternative form of] diabetes despite her very young age of onset and the presence of PAA, but this is still hard to determine completely.” Her T1D_GRS_AFR was 1.101 (<5^th^ percentile for T1D in MGBB). Her T1D_GRS_EUR was 8.13 (<5^th^ percentile for T1D in UKBB). Both T1D polygenic scores were low, suggesting that this is not classic T1D and may explain the prolonged duration of diabetes without the need to restart insulin.

## Discussion

Making an accurate diagnosis is critical in pediatric diabetes care. Yet, clinicians still struggle to do so consistently. Additional tools could help clinicians reach the correct diagnosis more often. A genetic approach, which has not been widely leveraged to this point, could yield significant power. Here, we demonstrate multiple scenarios where T1D polygenic scores could have immediate clinical utility to improve provider diagnostic accuracy.

Previous studies have suggested sufficient differentiating capacity of T1D polygenic scores for separating T1D from not-T1D, with an AUC around 0.90 [20, 18]. In this study, T1D polygenic scores by themselves also had an AUC 0.89 overall across individuals of all ancestries. This discriminating capacity, when combined with other measures, could be helpful to clinicians. The sensitivity or specificity of the test could be altered by adopting different cutoff points depending on the clinical scenario.

Both T1D and T2D are caused by a combination of environmental and genetic factors. While a genetic test by itself does not capture environmental risk factors, it could serve as an adjunct to other current tools for classifying diabetes diagnosis. Even though these tools may not seem easily implementable given non-immediate access to genetic data, genetic information is becoming more readily available and should become more widespread in the coming years.

The next step for utilizing T1D polygenic scores in clinic will involve developing a pipeline to return these results to patients and providers and ensure they are readily interpretable [31]. There are already multiple studies where T1D polygenic scores are being returned to patients to indicate risk for T1D. The Electronic Medical Records and Genomics (eMERGE) Network is returning T1D polygenic scores to delineate risk of T1D to 25,000 diverse adults and children as part of a clinical study [32]. T1D polygenic score return is also occurring in the Early Check study to families of infants [33].

A new test must be interpreted correctly to not confuse the diagnostic picture. As evident above, these scores are not perfect, with some individuals with a T1D polygenic score below the optimal point for distinguishing T1D from not-T1D still displaying a classic T1D picture. Additionally, an individual with MELAS had a T1D polygenic score above the optimal point for differentiating T1D from not-T1D. Had the T1D polygenic score been the only test utilized for diagnostic screening, this would have missed MELAS. Therefore, in cases where a presentation is atypical, even with a high T1D polygenic score, genetic testing should still be considered. As in the current diagnostic paradigm, it is recommended to send PAAs before genetic testing for MODY, as MODY is highly unlikely with PAA-positivity [34]. Therefore, a T1D polygenic score may be helpful as an adjunctive test when PAA screening is not informative or if questions remain after PAA testing. Even with autoantibodies, a low T1D polygenic score may suggest monogenic autoimmunity or an autoimmune polyglandular syndrome with a different genetic architecture than classical isolated T1D [23] (which was unable to be determined with the genetic data available).

Another key to clinical implementation of these scores is to ensure that they do not exacerbate health care disparities [35]. One outstanding question remains how to implement these scores across different ethnicities and ancestries. These scores were originally developed in European and African ancestries. Here, even with limited numbers, we show that these scores can discriminate between T1D and not-T1D, even in ancestries where they were not originally developed. However, the median score was lower for T1D_GRS_EUR in non-European ancestries than European ancestries. Additional studies are now needed, with larger pediatric cohort sizes, to determine optimal thresholds for differentiating T1D and not-T1D in different ancestries. Additional work continues to be needed to optimize the use of polygenic scores in multi-ancestry populations.

Strengths of this study include the computation of T1D polygenic scores in a clinical setting pediatric cohort, with a diversity of presentations and diagnoses, that will mirror situations where these scores would be used in clinical practice. Further, access to comprehensive medical information previously obtained allowed comprehensive investigation to verify diabetes classification and assess other parameters that may not be practical in large biobanks. Weaknesses include that samples did not have whole genome sequencing, allowing for implementation and testing of other polygenic scores or further investigation to search for monogenic causes of diabetes. Additionally, other clinical lab tests, like PAA, were not available for all patients. If these were present, the combination of T1D polygenic scores, PAA, and other clinical measures could be tested together to assess diagnostic accuracy, which will ultimately be the paradigm used in clinic.

## Conclusions

T1D polygenic scores have significant predictive capacity that could greatly augment current tools for correctly classifying pediatric diabetes. While T1D polygenic scores are already being returned to delineate the risk of developing diabetes in some studies, here we illustrate their potential utility for improving diabetes diagnosis in a real-world clinical setting. New research is now needed to optimize a pipeline for clinical implementation.

## Statements

## Acknowledgement

Portions of this manuscript were presented at the 2024 Pediatric Endocrine Society (PES) Annual Meeting. Dr. Raymond Kreienkamp acknowledges data support from the PrecisionLink Biobank for Health Discovery at Boston Children’s Hospital. We also acknowledge all the patients that participated in the PrecisionLink Biobank for Health Discovery at Boston Children’s Hospital.

## Statement of Ethics

The PLB initiative is approved by the BCH Institutional Review Board (protocol number - P00000159). This project (Genetic and clinical features of diabetes and metabolic conditions) is approved by the BCH Institutional Review Board (protocol number - P00044203). All individuals participating in PLB provided written informed consent (and assent, where appropriate) to do so.

## Author Contributions

RJK and MSU were involved in the conception of the project design. RJK, AJD, and AHC performed or contributed to analyses for the manuscript. EMB and JF were involved in procure discussion and reviewed the final version of the manuscript, as well as approved the final version for publication.

## Data Availability Statement

The data that support the findings of this study are available from Boston Children’s Hospital PrecisionLink Biobank. Restrictions apply to the availability of these data, which were used under license for this study. Data are available from the author(s) with the permission of Boston Children’s Hospital PrecisionLink Biobank.

